# Analytical Validation of an ELISA assay for Maternal Autoantibody Related Autism

**DOI:** 10.64898/2026.02.25.26347095

**Authors:** Mags Mcinerney, Beth Hurley, Jessica Barkow, Katherine Menning, Justin Nicolace, Joseph Schauer, Judy Van de Water, E. Robert Wassman

## Abstract

**Background:** The influence of genetic and environmental factors, especially during early development, is critical in the pathogenesis of autism. Maternal autoantibodies that recognize specific fetal brain proteins can be strong predictors of autism risk. These antibodies cross the placenta and bind to their target antigens, which play critical roles in neurodevelopment, thereby increasing autism risk. This etiologically defined subtype is now referred to as Maternal Autoantibody-Related Autism (MARA). The newly developed MAR-Autism^TM^ test is an indirect multi-ELISA assay designed to detect specific combinations of these maternal antibodies, which strongly predicts increased autism risk.

**Objective:** Translation of the indirect ELISA assays for the eight relevant antibodies (LDH-A, LDH-B, GDA, STIP1, CRMP1, CRMP2, NSE and YBOX) from an academic laboratory to a clinical development laboratory for optimization and determination of the analytical performance of the individual antibody assays.

**Methods:** Feasibility assays were transferred from the academic laboratory and their performance confirmed prior to optimization of all steps from target protein production to preliminary threshold determination. Validation to rigorous standards was conducted. The ELISAs are qualitative assays using an internal continuous response and a cutoff to define positivity and negativity for each analyte. Analytical performance metrics of linearity, sensitivity, specificity, precision, and stability were determined by standard testing methodologies.

**Results:** The optimized ELISAs all performed at acceptable standards for analytical performance. All of the assays except one were demonstrated to be linear upon dilution with buffer and with non-reactive plasma, however, recovery was overestimated with buffer diluent. The precision profile results demonstrated that the Lower Limit of Quantification (LOQ) was greater than the Limit of Detection (LOD) and below the preliminary thresholds determined from a general population cohort distribution. Precision studies showed coefficients of variation less than 15% with two minor exceptions. Common interfering substances, apart from whole human IgG, did not affect assay performance. The microtiter assay plates were stable for at least 6 months without significant drift.

**Conclusion:** Overall, the individual antibody assays demonstrated high sensitivity, specificity, and robustness sufficient to enable extension to clinical validation. These assays enable evaluation of specific antibody combinations that were previously reported to strongly and specifically correlate with autism risk, particularly in settings of suspected diagnosis or in families with an older sibling with a confirmed autism diagnosis.

## Introduction

Autism is a neurodevelopmental condition characterized by impairments in social interaction and communication, along with restricted or repetitive patterns of thought and behavior.^1^ Early developmental risk for autism is shaped not only by genetics, but also by environmental factors, including dysregulation of the maternal immune system during gestation.^2–4^ Maternal autoantibodies reactive to fetal brain proteins were demonstrated to be strong potential predictors of autism risk in a subset of cases in research studies in the Van de Water lab at UC Davis/MIND Institute.^4–10^ Reactivity to eight proteins highly expressed in the developing brain occurring in specific combinations, referred to as Maternal Autoantibody-Related Autism (MARA), is associated with an increased likelihood of autism in the offspring. ^4–12^ During early brain development in pregnancy, when the blood brain barrier (BBB) is more permissive, thus allowing maternal autoantibodies can cross the placenta and access the developing fetal brain.^13–14^

Biomarkers are increasingly critical to the diagnosis and management of patients in clinical research, drug development, and especially in the clinic. However, the lack of mechanistic links to the pathology underlying autism has heretofore been limiting. Further, current clinical diagnosis, while accurate, rests on detailed clinical-psychological evaluations, resulting in long diagnostic odysseys and relatively late definitive diagnosis, often when the child is 3-5 years old.^15^ Van de Water’s research represents a clear potential for these as predictive biomarkers of autism, based on discrete combinations of maternal autoantibodies that highly correlated with gold standard clinical diagnosis of autism in offspring.^4–12^ These findings, along with animal model studies demonstrating autism-relevant outcomes following gestational exposure to the same autoantibodies — either by passive transfer or active antibody production by the dams — support the likelihood that MARA are not merely associated biomarkers, but pathogenic agents.^1,16–23^ The identification of mechanistic biomarkers makes the rigorous translation of these findings to the clinic an important and valuable path to earlier diagnosis and potential therapeutic/preventative intervention in a subset of autistic individuals.

Enzyme-linked immunosorbent assays (ELISAs) used in research studies are powerful and practical tools for measurement of low-abundance autoantibodies. However, methodological variations in ELISA assays may introduce both systematic and random errors as well as poor clinical quality. The first step in clinical translation of these important findings is rigorous method validation to control assay performance suitable for its intended use.^24^ A predefined validation protocol with established acceptance criteria for each parameter was used to validate this set of qualitative indirect ELISA autoantibody assays as a laboratory-developed test. The assays detect previously identified maternal autoantibodies to key proteins highly expressed in the developing brain: anti-CRMP2, anti-GDA, anti-CRMP1, anti-STP1, anti-NSE, anti-LDHA, anti-LDHB, and anti-YBOX.^25–35^ Accordingly, this paper presents the validation protocols and analytical performance evaluations that establish the suitability and reliability of the MAR-Autism^TM^ test (Marabio, Inc., Salt Lake City, Utah) for its intended clinical use.

## Materials and Methods

### Specimen Sources

Blood samples used to evaluate the analytical performance of the individual ELISAs in the MAR-Autism^TM^ test were sourced from the San Diego Blood bank under IRB oversight. Samples used to verify the preliminary cutoff for each assay were obtained from the Van de Water laboratory at UC Davis.^11^ These samples had been classified as MAR-Autism positive or negative using the original feasibility assay developed at UC Davis.

Samples used to validate the CLIA Laboratory-Developed Test (LDT) were obtained from the Van de Water laboratory at UC Davis. Two sets of samples were used: 1) samples classified as MAR-Autism positive or negative using the original feasibility assay developed at UC Davis^11^ and 2) clinical samples from the CHARGE dataset classified as antibody positive or negative based on the preliminary cutoffs at the development lab^5–10,12^.

### Protein Production

Recombinant His-tagged proteins for multiple MAR-Autism ELISAs were produced using a baculovirus expression system in *Spodoptera frugiperda* (Sf9) or *Trichoplusia ni* (Tn5) derived insect cells. Codon-optimized genes were chemically synthesized, cloned into baculovirus transfer plasmids, and used to generate recombinant virus. Following infection and expression, cells were harvested, lysed, and the proteins purified by nickel-affinity chromatography, desalted, and sterile-filtered. Protein concentration was quantified using the bicinchoninic acid (BCA) protein assay. Proteins were then aliquoted, frozen, and shipped on dry ice for use in coating 96-well ELISA plates.

### ELISA Plate Preparation Procedure

The recombinant antigens (proteins) were diluted with standard coating buffer to appropriate concentrations. 100uL of these mixtures was added to each well of a 96 well plate, the plate sealed and incubated overnight at 4°C. Plates were washed on a plate washer with wash buffer. After the final wash, excess wash buffer was removed, and 200ul/well of blocking buffer per well was added and incubated in the covered plate for 1 hour at room temperature. Plates were stored at 2-8 °C until use.

### MAR-Autism^TM^ ELISA Procedure Methodology

The tests were performed as indirect ELISAs. Diluted acid–citrate–dextrose (ACD) plasma samples, calibrator, and controls were incubated in microwells coated with purified recombinant proteins. A single point calibration method was used along with a reagent blank control. Either a 1:250 or 1:500 dilution of the calibrator, controls, and plasma samples in sample diluent was prepared. 100 uL of diluted calibrator, controls, and plasma samples were added to the appropriate microwells in duplicate (including the reagent blank) and the covered plate was incubated at room temperature for 90 minutes. Following incubation, the plate was washed four times with wash solution using an automated plate washer without allowing the wells to dry out at any time. After the removal of unbound proteins by washing, 100uL of a previously prepared 1:10,000 dilution of a commercial Goat anti-Human IgG horseradish peroxidase (HRP)-Conjugated Antibody (Seracare, Milford, MA) was added to each well to form complexes with the target protein bound antibodies. The plate was covered again with a plate sealer and incubated for 60 minutes at room temperature. After incubation with the goat-anti-human IgG, plates were washed again in the same manner four times. Following another washing step, the bound enzyme-antibody conjugate was assayed by the addition of a one-component substrate solution (100uL) containing tetramethylbenzidine (TMB) and hydrogen peroxide (H_2_O_2_) as the chromogenic substrate and incubated for 30 minutes at room temperature. Stopping solution (100uL of 0.36 N sulfuric acid) was then added to each well in the same order and at the same rate as the substrate was added to stop the enzyme reaction. In positive samples, blue color develops in the wells at an intensity proportional to the concentration of protein-specific antibodies. Results were obtained by reading the OD of each well at 450 nm in a spectrophotometer within 5 minutes of adding the stopping solution, using a reference of 595 nm. The %CV for duplicate wells of calibrator, controls and samples with a mean OD >0.100 should be within ±10%. For calibrator, control, or samples with mean OD ≤ 0.100, the difference between duplicate wells should be ≤0.050 OD. Readings of the Calibrator OD should preferably be ≥ 0.250 OD to assure the assay is functioning properly, and the reagent blank should preferably be ≤ 0.150 OD. The low control should recover less than the calibrator (< 100 ELISA Units (EU)). And the high control should recover greater than the calibrator (> 100 EU).

### Result Calculation and Quality Control

Studies were carried out in the development lab using calibrated equipment with reagents and materials, including samples from UC Davis and the San Diego Blood Bank, which were procured and stored according to established procurement policies and quality system protocols. Data and reports were generated and stored according to the development lab’s quality system. All studies were run using a single calibrator close to the estimated cutoff plus one positive and one negative control. The calibrator value was normalized to 100 EU. A conversion Factor (CF) was determined for each assay run by dividing the calibrator value (100 EU) by the calibrator mean OD. Control and sample ODs from the same assay were multiplied by the CF to obtain an antibody concentration value expressed in EU units.

### Overview of Pathway from Feasibility to Analytical Validation

Eight qualitative indirect ELISAs to detect the presence of antibodies in maternal plasma specific to CRMP2, GDA, CRMP1, STP1, NSE, LDHA, LDHB, and YBOX —previously associated with autism by the Van de Water laboratory^4–11^ — were translated into reproducible LDT assays to ensure that the performance characteristics of the method were suitable and reliable for the intended clinical testing applications. Validation was completed to College of American Pathologists (CAP) requirements using Clinical and Laboratory Standards Institute (CLSI) guidelines as appropriate to help guide study design.

This process entailed four phases: 1) technology transfer; 2) assay optimization; 3) analytical performance evaluation and establishment & verification of preliminary cutoffs and 4) clinical validation (results to be published in a separate paper).

1. Technology Transfer: Feasibility of the ELISAs was established in the Van de Water lab at UC Davis, as described in the literature. ^4–11^ During the technology transfer phase, eight assay protocols from UC Davis were transferred unaltered into the development lab (Corgenix, Inc., Boulder, CO) and sufficient testing was conducted to establish equivalence between the two locations through sample bridging studies.
2. Optimization: Reagents and parameters were then optimized for best assay performance in a clinical lab setting. The key factors included in the assay parameter optimization studies were: 1) optimization of the protein coating concentration; 2) optimization of the sample dilution and dilutional linearity studies; 3) optimization of the secondary antibody (IgG-HRP conjugate) dilution; 4) optimization of plate coat buffer reagent exchange and accelerated stability studies; and 5) bridging studies between independent lots of proteins to confirm assay parameters and lot-to-lot variability of proteins. Bridging studies with three independently produced protein lots were conducted on pre-pilot reagent lots.
3. Analytical Performance Evaluation and Establishment and Verification of Preliminary Cutoffs: Optimization was followed by production of three independent pilot lots to evaluate the performance characteristics of the assays prior to analytical validation. The designs used to evaluate each of the MARA autoantibody ELISAs were identical, were based on CLSI guidelines where applicable,^36–41^ and included linearity, precision, analytical sensitivity [Limit of Blank (LOB), Limit of Detection (LOD), Limit of Quantitation (LOQ)], analytical specificity (interference) and stability (shelf-life). Once the performance of the ELISAs was determined to be acceptable, a single lot of plates was used to establish preliminary cutoffs for each assay using a set of 200 samples from the San Diego Blood Bank (San Diego, CA). The cutoffs were established using the 97.5^th^ percentile of the normal distribution of samples.
4. Analytical Validation: Analytical validation studies were performed in two independent Clinical Laboratory Improvements Act (CLIA) laboratories following the College of American Pathologists’ (CAP) checklists relevant for performing similar assays in CAP certified laboratories.^42,43^ This analytical validation testing included: linearity (diluent & negative plasma sample), precision (within-run and between-run), accuracy (using previously tested clinical samples), analytical sensitivity (LOD, LOB, LOQ), analytical specificity (interference), sample shipping and storage, plate stability and lot-to-lot comparison.

### Study Design: Performance Evaluation Studies Linearity & Recovery

Linearity was determined with one protein/plate lot for each ELISA using a high OD and a low OD ACD plasma sample. The high sample was diluted in a 5-point serial dilution with both a low OD sample and a non-plasma diluent. Samples from three separate dilution preparations were assayed in duplicate and the mean, standard deviation and %CV for each sample were calculated. Recovered values were compared to expected values using linear regression. The guideline targets used to assess the preliminary performance were a slope of ≥ 0.90 and recovery of +/- 10%.

### Analytical Sensitivity (LOB/LOD/LOQ)

Evaluation based on guidance from CLSI EP17-A2.^39^ Analytical sensitivity refers to the lowest concentration of a substance that an assay is capable of measuring accurately in a sample. It is determined under controlled laboratory settings and focuses mainly on evaluating the assay’s technical performance. Analytical sensitivity, i.e., limits of detection and quantitation, were assessed using one lot of protein for each antigen. LOB was determined as the mean of 40 determinations using a pool of two negative samples. The LOD is based on the mean of 20 determinations from a low-level sample (one high and one negative ACD plasma sample mixed to give five low level samples) with one run per day for two days and ten replicates per run. It is distinguished from the LOB based on the upper (LOB) and lower (LOD) 95% confidence intervals. A precision profile was prepared to determine the LOQ based on the mean of 20 determinations from one of the precision profile levels (pool of negative and positive ACD plasma), assayed in ten measurements on each of two days. The LOQ was defined as the preparation with the lowest %CV level from the precision profile results where LOQ ≥ LOD. The mean, standard deviation, %CV, and confidence interval (CI) for the LOB, LOD, and LOQ for each sample were calculated.

### Precision

Evaluation based on guidance from CLSI EP05-A3.^36^ Precision (repeatability and reproducibility) was assessed by a 5 x 5 x 3 matrix analysis, i.e., five replicates (mean of duplicates) of all three protein/plate lots, run once daily over five days on four ACD plasma samples spaced across the following range: 1) negative (lowest OD available), 2) high (highest OD available); plus two admixtures of these giving 3) low, and 4) moderate test points. Outliers were identified using the Grubbs test.^44^ Repeatability (within-run) and reproducibility (between-day, within-lot, between lot) %CVs were calculated. Inter-operator precision was not evaluated in these studies.

### Analytical Specificity (Interference)

Evaluation based on guidance from CLSI EP07-A3.^41^ For initial interference evaluations, the following substances (concentrations) were spiked into the samples (one lot of protein per antigen; one positive and one negative ACD sample pool) prior to assay: hemoglobin (1000mg/dL), Intralipid (1000mg/dL), bilirubin (conjugate and unconjugated) (40mg/dL), rheumatoid factor (RF) (500 IU/mL), IgG (4800mg/dL) and anti-cardiolipin IgG (a systemic lupus erythematosus [SLE]–positive antibody). IgG was tested in CRMP2 and GDA only. Testing was discontinued due to interference at all levels.

Three known SLE-positive samples were then compared to the negative control, and the preliminary cutoff for each ELISA was applied in subsequent testing to estimate this potential for interference. A total of 26 replicates of positives and negatives with and without interferents were tested (except SLE samples). Mean, standard deviation and %CV were calculated for each condition and interferent was compared to control in each case and deemed non-interfering if within ±20%. If this criterion was not met, dose response testing was performed to determine the concentration of interferent meeting the criteria.

### Stability (Plate Shelf-life at 4°C)

Each protein was assessed across three plate lots with four ACD plasma samples spanning the range from negative (lowest OD available) to high (highest OD available), plus two admixtures at low and moderate level ODs. Two replicates (independent dilutions) were tested in duplicate at multiple timepoints up to six months. Means and standard deviations derived from precision testing was used as a guideline for the limit of allowable drift. Assessment of stability was made on the basis of visual examination using the most conservative approach.

### Establishment and Verification of Preliminary Cutoffs

1. Cutoff Establishment: Preliminary cutoffs were established for each assay using a set of ACD plasma samples collected from 200 female donors at the San Diego Blood Bank, meeting their health screening criteria for blood donation. Assay results were used to define the reference interval in the general population and preliminary thresholds were generated for each ELISA in the development lab. The results were analyzed for normal distribution and the following sets of thresholds for positivity were suggested: mean+2SD, mean+3SD and 97.5^th^ percentile.
2. Cutoff Verification: Qualified donor samples (N = 24 to 29) were assigned as positive or negative according to the UC Davis feasibility ELISAs and tested on the optimized ELISAs. Cutoffs were selected based on the thresholds that lead to the greatest separation of positive and negative samples in the verification set. The mean +3SD, 97.5^th^ percentile and Youden Index (YI) were applied to the verification data set to determine the %Agreement for each ELISA to complete cutoff selection.

### CLIA Validation

Once the thresholds for positivity were determined the test was transferred into two CLIA and CAP certified clinical laboratories and validated to demonstrate that the previously evaluated performance metrics could be replicated prior to clinical use (Corgenix Clinical Laboratories, Bolder CO and Invitrox, Raleigh NC). The protocols and study designs adhered to CLIA lab procedures for validation of assays, met CAP requirements, and were approved by each laboratory’s Medical Directors. The studies were carried out under the CLIA labs’ quality systems using appropriately calibrated equipment and trained personnel.

The following studies were performed in accordance with the lab’s procedures: linearity (diluent & negative sample), precision (within-run and between run), accuracy (versus results for clinical samples), reportable range (reference interval), analytical specificity (interference), analytical sensitivity (LOD, LOB, LOQ), sample shipping and storage, plate stability and lot-to-lot comparison.

## RESULTS

All eight individual autoantibody assays demonstrated satisfactory performance during the performance evaluation phase. Table 1 summarizes the results of the performance evaluation of each ELISA for precision, sensitivity, specificity, linearity, and assay plate stability.

**Table 1:** Summary of Performance Evaluation Data.

| ELISA | Precision [%CV] *<br>(Low Level Sample) | Sensitivity [LOQ] @<br>(LOQ < Cutoff) | Specificity^ | Linearity [R<sup>2</sup>] #<br>Buffer/Plasma | Stability |
| --- | --- | --- | --- | --- | --- |
| Anti-CRMP2 | 15.0% (184 EU) | 59 EU (Yes) | Acceptable | 0.99/0.99 | 6 months |
| Anti-CRMP1 | 9.7% (182 EU) | 57 EU (Yes) | Acceptable | 0.89/0.89 | 6 months |
| Anti-GDA | 18.9% (133 EU) | 41 EU (Yes) | Acceptable | 0.99/0.99 | 6 months |
| Anti-STIP1 | 10.6% (143 EU) | 61 EU (Yes) | Acceptable | 0.99/0.99 | 6 months |
| Anti-NSE | 17.3% (220 EU) | 33 EU (Yes) | Acceptable | 0.99/0.99 | 6 months |
| Anti-YBOX | 7.1% (181 EU) | 57 EU (Yes) | Acceptable | 0.95/0.98 | 6 months |
| Anti-LDHA | 10.5% (136 EU) | 48 EU (Yes) | Acceptable | 0.98/0.99 | 6 months |
| Anti-LDHB | 11.2% (147 EU) | 57 EU (Yes) | Acceptable | 0.99/0.99 | 6 months |

\*Precision (Total reproducibility) for 3 lots/5 days/2 replicates determined close to the estimated cutoff
@Sensitivity/LOQ is the lowest level at which performance is acceptable
^Specificity with standard interferants, hemoglobin, bilirubin, lipid, rheumatoid factor

**Figure 1.**
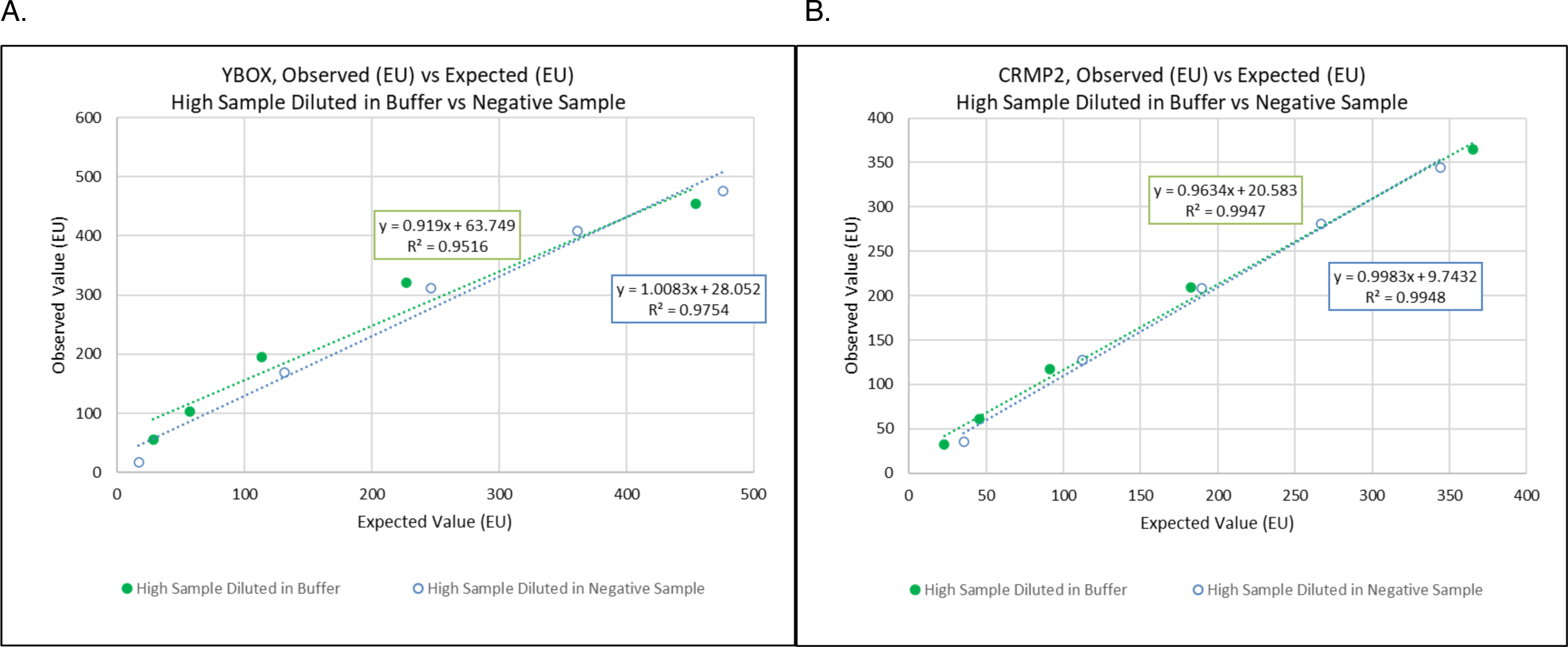
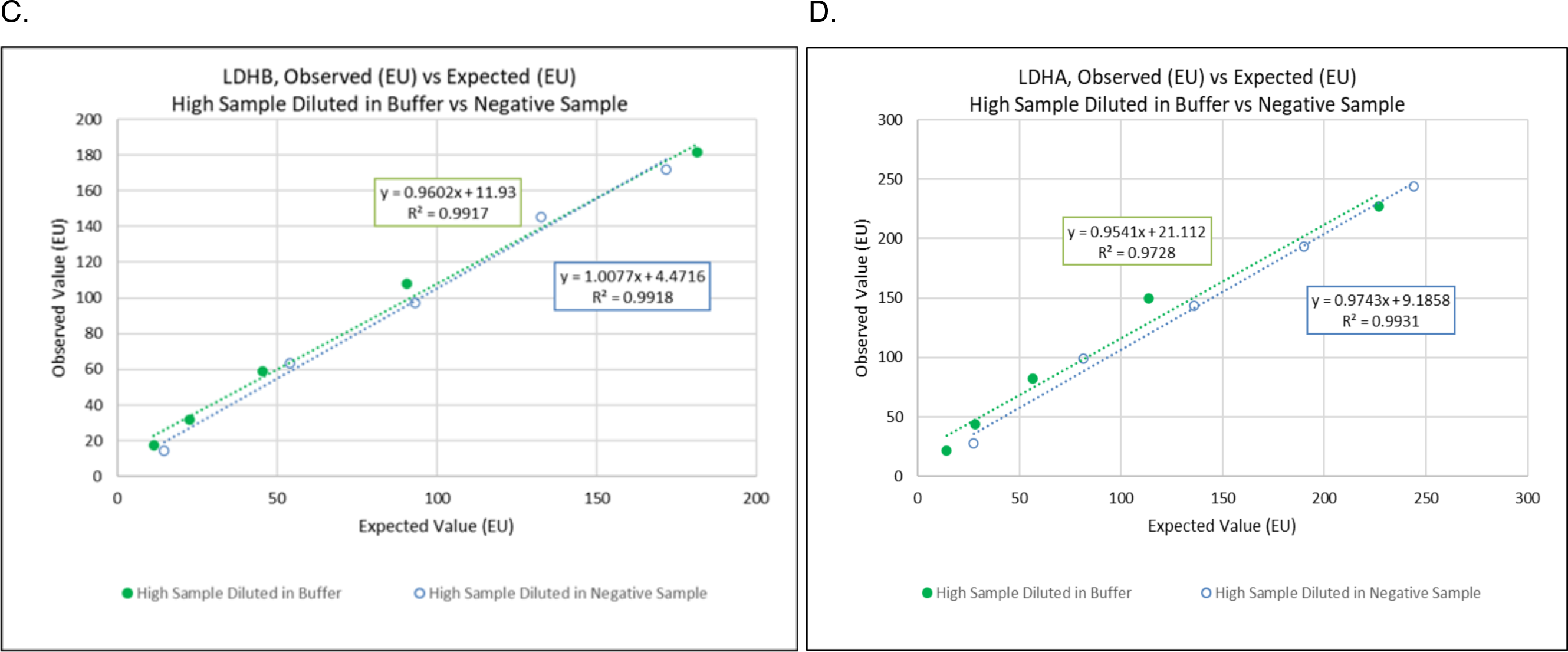
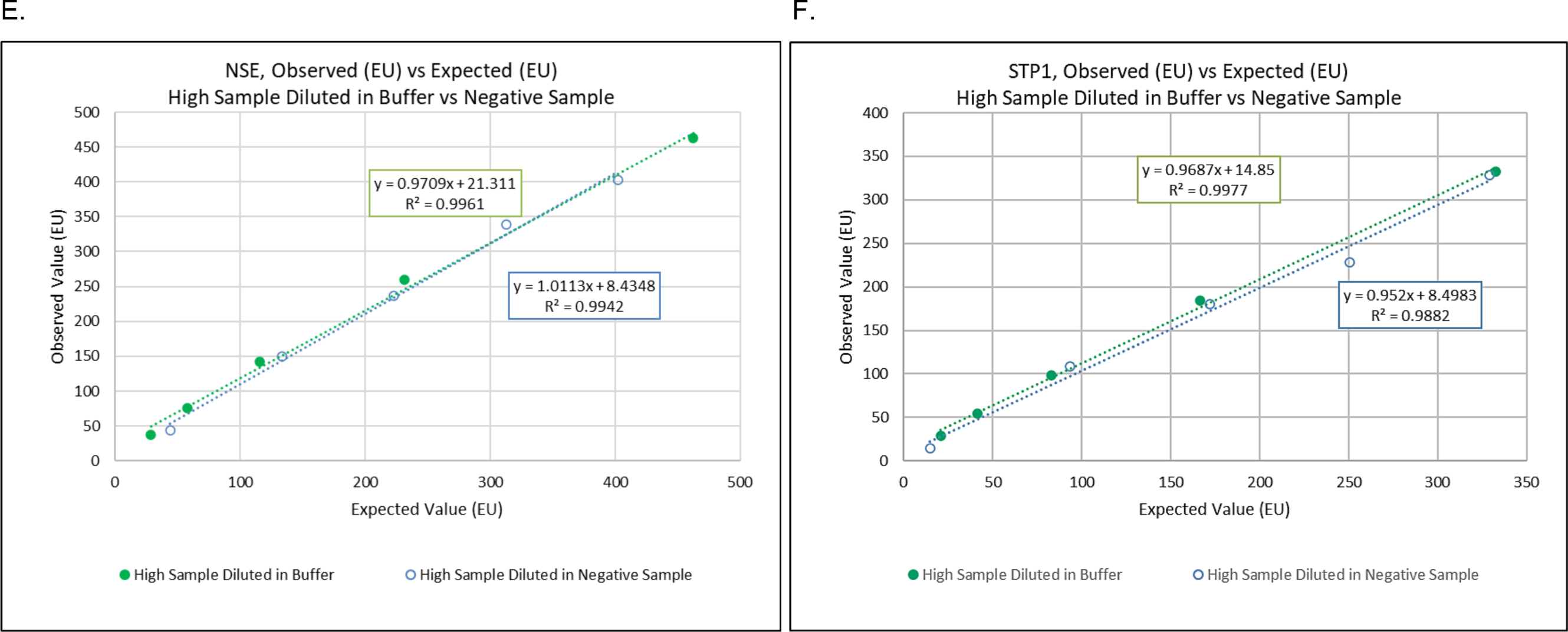
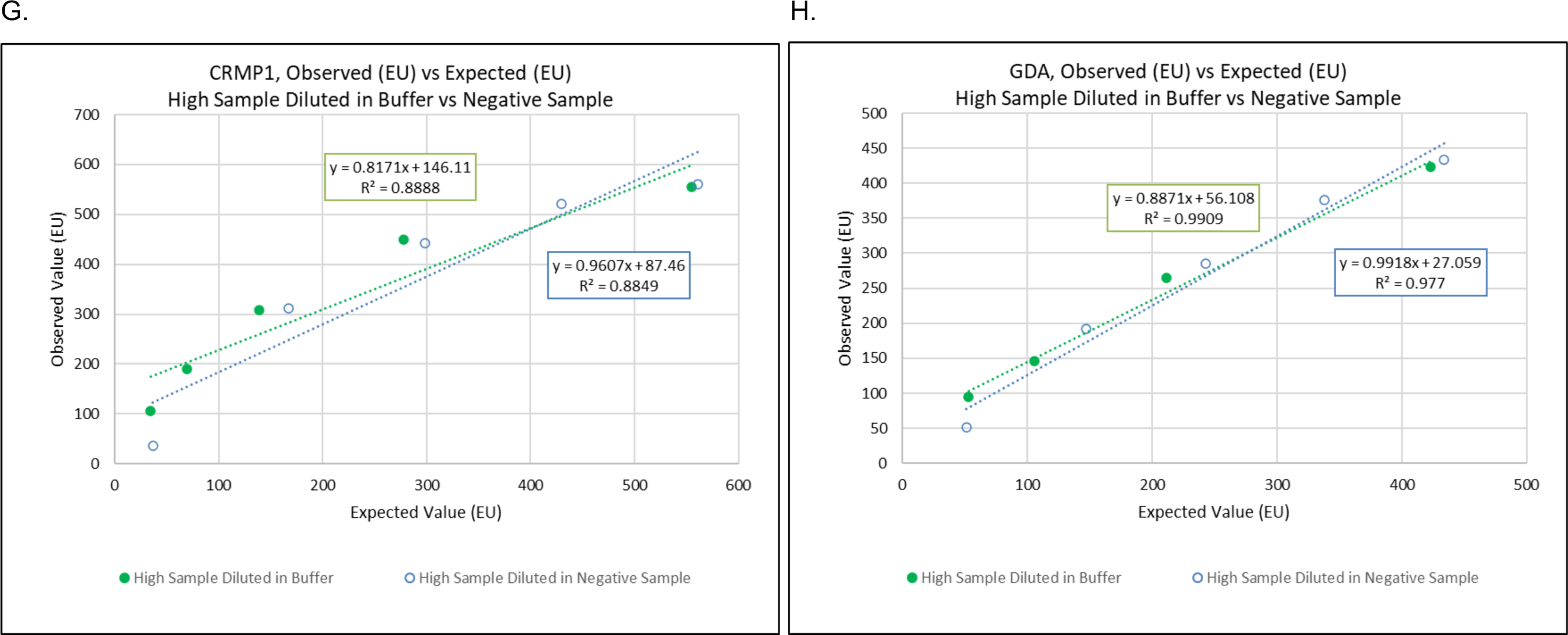
**A-H.** Linearity results for each ELISA for the sample with the highest OD diluted in a) non-plasma diluent and b) low OD ACD plasma. Abbreviation: EU, ELISA units

### Linearity

Linear regression (R^2^) was 0.99 for both diluents for six of the eight ELISAs, while anti-YBOX was 0.95/0.98 and anti-CRMP1 was 0.89/0.89 (buffer/plasma), indicating non-linearity in this assay. Dilutions using non-plasma diluent demonstrated significant overestimation of antibody levels for all assays indicating dilution of samples above the capability of the spectrophotometer should be diluted with a low OD plasma rather than non-plasma diluent. Overall, the linearity of the assays was determined to be acceptable.

### Sensitivity

Limit of Blank (LOB), Limit of Detection (LOD), and Lower Limit of Quantification (LOQ) were determined for each antibody assay (Table 2).

**Table 2.** LOB, LOD, and LOQ for each antigen.

| Antigen | LOB (EU) | LOD (EU) | LOQ (EU) |
| --- | --- | --- | --- |
| **LDHB** | 10.9 | 19.4 | 57.3 (3.7% CV) |
| **LDHA** | 26.6 | 38.6 | 47.6 (5.5% CV) |
| **YBOX** | 18.8 | 38.8 | 57.2 (6.8% CV) |
| **STP1** | 19.0 | 26.6 | 32.9 (8.4%) |
| **NSE** | 38.1 | 50.9 | 61.1 (10% CV) |
| **GDA** | 19.0 | 30.8 | 41.1 (5% CV) |
| **CRMP1** | 31.1 | 46.6 | 56.7 (6%) |
| **CRMP2** | 41.1 | 54.1 | 59.4 (5% CV) |

EU, ELISA units

The mean LOB results ranged from 10.9 EU to 41.1 EU and in each case the 95% upper limit of CI was below the 95% lower limit of the respective LOD’s means which ranged from 19.4 EU to 54.1. The LOQs ranged from 32.9 EU to 61.1 EU with all CVs ≤10%. For each assay the LOQ was > LOD, and all LOQs were below the subsequently determined preliminary thresholds.

### Precision

Tables 3 to 10 describe the Precision (repeatability and reproducibility) of each ELISA tested across four levels (1=negative, 2= low OD, 3=moderate OD and 4=high OD). Reproducibility was tested across three plate/protein lots for each ELISA.

**Table 3.** YBOX Summary of Precision.

| *Level* | *Mean (EU)* | *n* | *Repeatability* |  | *Between Day* |  | *Within Lot* |  | *Between Lot* |  | *Total Reproducibility* |  |
| --- | --- | --- | --- | --- | --- | --- | --- | --- | --- | --- | --- | --- |
|  |  |  | *SD* | *%CV* | *SD* | *%CV* | *SD* | *%CV* | *SD* | *%CV* | *SD* | *%CV* |
| 1 | 18.0 | 75 | 1.3 | 7.3% | 2.4 | 13.4% | 2.8 | 15.3% | 0.1 | 0.6% | 2.8 | 15.3% |
| 2 | 180.5 | 75 | 6.2 | 3.4% | 12.0 | 6.6% | 13.5 | 7.5% | 0.0 | 0% | 12.9 | 7.1% |
| 3 | 290.7 | 75 | 10.3 | 3.6% | 22.4 | 7.7% | 24.7 | 8.5% | 0.0 | 0% | 22.7 | 7.8% |
| 4 | 575.9 | 75 | 19.6 | 3.4% | 62.7 | 10.9% | 65.7 | 11.4% | 0.0 | 0% | 59.8 | 10.4% |

EU, ELISA units; SD, Standard deviation; CV, coefficient of variation

**Table 4.** LDHB Summary of Precision.

| *Level* | *Mean (EU)* | *n* | *Repeatability* |  | *Between Day* |  | *Within Lot* |  | *Between Lot* |  | *Total Reproducibility* |  |
| --- | --- | --- | --- | --- | --- | --- | --- | --- | --- | --- | --- | --- |
|  |  |  | *SD* | *%CV* | *SD* | *%CV* | *SD* | *%CV* | *SD* | *%CV* | *SD* | *%CV* |
| 1 | 8.7 | 75 | 1.8 | 21.1% | 2.1 | 24.2% | 2.8 | 32.1% | 0.2 | 1.9% | 2.8 | 32.2% |
| 2 | 147.2 | 75 | 7.6 | 5.2% | 16.3 | 11.1% | 18.0 | 12.2% | 0.0 | 0.0% | 16.4 | 11.2% |
| 3 | 193.7 | 75 | 12.6 | 6.5% | 21.5 | 11.1% | 24.9 | 12.9% | 0.0 | 0.0% | 23.0 | 11.9% |
| 4 | 233.7 | 75 | 11.5 | 4.9% | 31.4 | 13.4% | 33.5 | 14.3% | 0.0 | 0.0% | 30.6 | 13.1% |

EU, ELISA units; SD, Standard deviation; CV, coefficient of variation

**Table 5.**
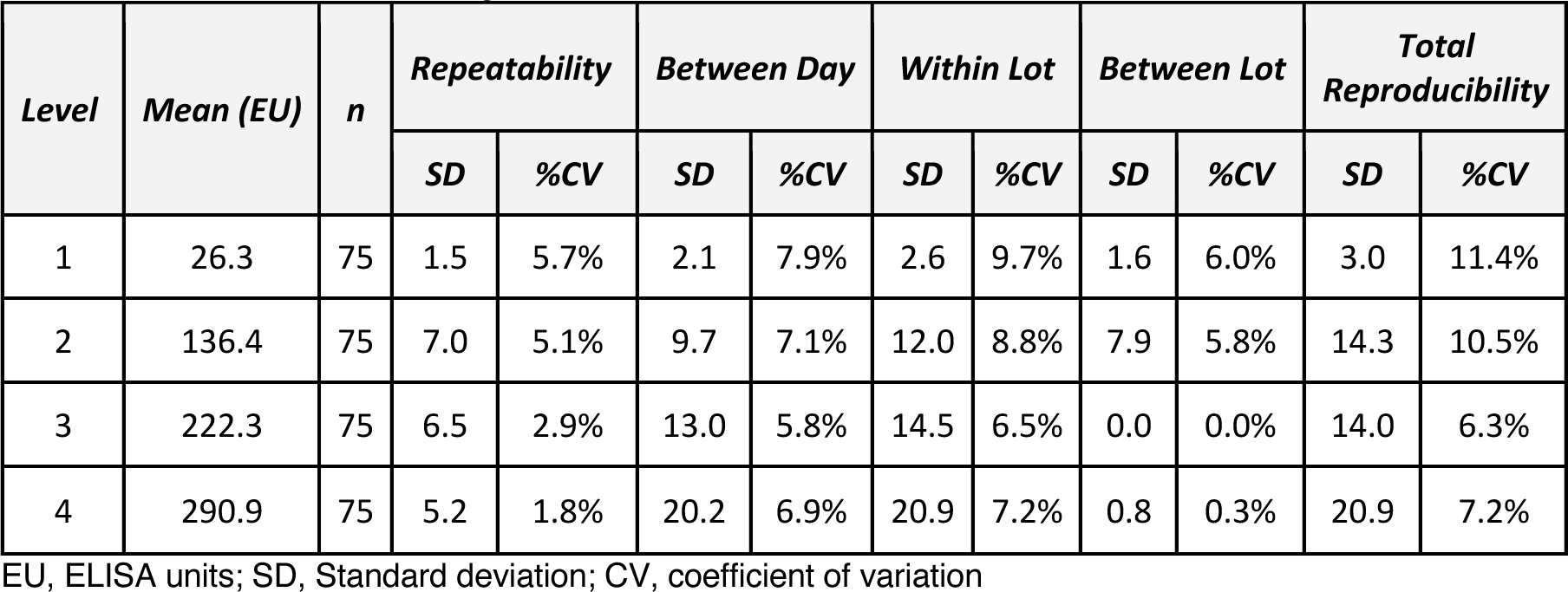
LDHA Summary of Precision.

**Table 6.** STIP1 Summary of Precision.

| *Level* | *Mean (EU)* | *n* | *Repeatability* |  | *Between Day* |  | *Within Lot* |  | *Between Lot* |  | *Total Reproducibility* |  |
| --- | --- | --- | --- | --- | --- | --- | --- | --- | --- | --- | --- | --- |
|  |  |  | *SD* | *%CV* | *SD* | *%CV* | *SD* | *%CV* | *SD* | *%CV* | *SD* | *%CV* |
| 1 | 28.6 | 75 | 1.7 | 5.8% | 3.4 | 11.8% | 3.8 | 13.2% | 10.3 | 35.9% | 10.9 | 38.2% |
| 2 | 142.9 | 75 | 7.4 | 5.2% | 13.2 | 9.2% | 15.2 | 10.6% | 1.4 | 1.0% | 15.2 | 10.6% |
| 3 | 189.9 | 75 | 10.9 | 5.7% | 15.8 | 8.3% | 19.2 | 10.1% | 0.0 | 0.0% | 18.8 | 9.9% |
| 4 | 280.8 | 75 | 12.3 | 4.4% | 23.0 | 8.2% | 26.1 | 9.3% | 26.6 | 9.5% | 37.3 | 13.3% |

EU, ELISA units; SD, Standard deviation; CV, coefficient of variation

**Table 7.** CRMP1 Summary of Precision.

| *Level* | *Mean (EU)* | *n* | *Repeatability* |  | *Between Day* |  | *Within Lot* |  | *Between Lot* |  | *Total Reproducibility* |  |
| --- | --- | --- | --- | --- | --- | --- | --- | --- | --- | --- | --- | --- |
|  |  |  | *SD* | *%CV* | *SD* | *%CV* | *SD* | *%CV* | *SD* | *%CV* | *SD* | *%CV* |
| 1 | 24.0 | 75 | 1.8 | 7.3% | 2.7 | 11.3% | 3.2 | 13.5% | 0.0 | 0.0% | 3.0 | 12.4% |
| 2 | 181.5 | 75 | 6.0 | 3.3% | 17.8 | 9.8% | 18.8 | 10.4% | 0.0 | 0.0% | 17.5 | 9.7% |
| 3 | 311.3 | 75 | 15.7 | 5.1% | 53.0 | 17.0% | 55.3 | 17.8% | 0.0 | 0.0% | 51.2 | 16.4% |
| 4 | 589.7 | 75 | 11.3 | 1.9% | 100.1 | 17.0% | 100.8 | 17.1% | 0.0 | 0.0% | 91.7 | 15.6% |

EU, ELISA units; SD, Standard deviation; CV, coefficient of variation

**Table 8.** CRMP2 Summary of Precision.

| *Level* | *Mean (EU)* | *n* | *Repeatability* |  | *Between Day* |  | *Within Lot* |  | *Between Lot* |  | *Total Reproducibility* |  |
| --- | --- | --- | --- | --- | --- | --- | --- | --- | --- | --- | --- | --- |
|  |  |  | *SD* | *%CV* | *SD* | *%CV* | *SD* | *%CV* | *SD* | *%CV* | *SD* | *%CV* |
| 1 | 30.5 | 75 | 5.0 | 16.3% | 3.4 | 11.2% | 6.0 | 19.7% | 0.0 | 0.0% | 5.8 | 19.2% |
| 2 | 184.1 | 75 | 16.7 | 9.1% | 13.8 | 7.5% | 21.7 | 11.8% | 17.2 | 9.4% | 27.7 | 15.0% |
| 3 | 232.1 | 75 | 14.9 | 6.4% | 23.3 | 10.0% | 27.7 | 11.9% | 12.4 | 5.3% | 30.3 | 13.1% |
| 4 | 329.4 | 75 | 33.0 | 10.0% | 29.4 | 8.9% | 44.1 | 13.4% | 41.9 | 12.7% | 60.9 | 18.5% |

EU, ELISA units; SD, Standard deviation; CV, coefficient of variation

**Table 9.** GDA Summary of Precision.

| *Level* | *Mean (EU)* | *n* | *Repeatability* |  | *Between Day* |  | *Within Lot* |  | *Between Lot* |  | *Total Reproducibility* |  |
| --- | --- | --- | --- | --- | --- | --- | --- | --- | --- | --- | --- | --- |
|  |  |  | *SD* | *%CV* | *SD* | *%CV* | *SD* | *%CV* | *SD* | *%CV* | *SD* | *%CV* |
| 1 | 47.5 | 75 | 3.7 | 7.7% | 2.4 | 5.0% | 4.4 | 9.2% | 1.7 | 3.6% | 4.7 | 9.9% |
| 2 | 132.9 | 75 | 7.6 | 5.7% | 7.8 | 5.8% | 10.8 | 8.2% | 22.7 | 17.1% | 25.1 | 18.9% |
| 3 | 258.6 | 75 | 13.4 | 5.2% | 14.8 | 5.7% | 20.0 | 7.7% | 22.2 | 8.6% | 29.9 | 11.6% |
| 4 | 429.7 | 75 | 26.4 | 6.1% | 41.8 | 9.7% | 49.4 | 11.5% | 31.6 | 7.4% | 58.7 | 13.7% |

EU, ELISA units; SD, Standard deviation; CV, coefficient of variation

**Table 10.**
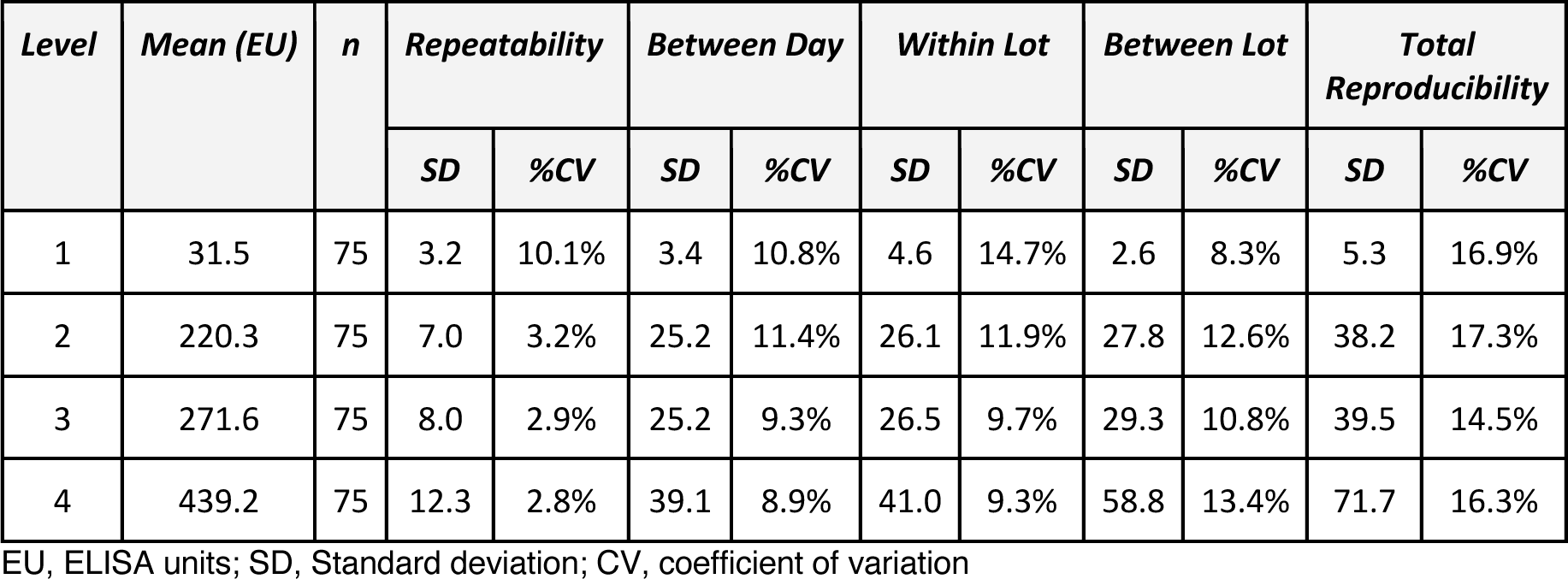
NSE Summary of Precision.

Targets for repeatability were 15% for Level 1 and 10% for Levels 2 – 4. The CVs ranged from 6.5% to 21.1% with 30/32 of the results being ≤ 10% and 15/32 being ≤ 5%. The two values above 10% were Level 1 CRMP1 (16.3%) and Level 1 YBOX (21.1%). Targets for within-lot repeatability were 20% for Level 1 and 15% for Levels 2 – 4. The CVs ranged from 1.9% to 32.1% with 31/32 of the results being ≤ 20% and 28/32 being ≤ 15%. The four values above 15% were Level 1 CRMP1 (19.7%), Level 1 LDHB (34.7%) and Level 3 &4 CRMP2 (17.8% & 17.1% respectively). The magnitude of the lot-to-lot %CV indicated that reproducibility depends on the ELISA process, indicating some potential variability in the protein lots and the plate coating process. This variability can be addressed going forward with ongoing process control. Overall, the precision/reproducibility of the assays was determined to be acceptable to proceed with further validation of the ELISAs.

#### Stability

(Plate Shelf-life at 4°C)

Real time and accelerated stability testing to date showed plates met stability requirements for all antibodies out to six months from date of preparation.

### Analytical Specificity (Interference)

No interference was observed at high concentrations of the common interference test substances, hemoglobin, intralipid, conjugated and unconjugated bilirubin, or rheumatoid factor, in either positive or negative plasma samples. IgG concentrations intended to simulate pathological levels were tested in anti-CRMP2 and anti-GDA ELISAs only. Testing was discontinued due to interference at all levels of IgG. Testing was substituted with three known SLE positive samples. Results were calculated following the generation and verification of the preliminary cutoff for each ELISA. EU values for each sample were assessed to determine whether it was above or below the preliminary cutoff. Results from the three samples are presented in Table 11 below.

**Table 11.** Anti-cardiolipin IgG (SLE) interference results.

| **Antigen** | **SLE Sample 1**<br>**(EU)** | **SLE Sample 2**<br>**(EU)** | **SLE Sample 3**<br>**(EU)** | **Preliminary**<br>**Cutoff (EU)** | **EU Sample**<br>**< EU Cutoff?** |
| --- | --- | --- | --- | --- | --- |
| **LDHB** | 39.2 | 47.0 | 51.7 | 64.4 | Yes |
| **LDHA** | 71.0 | 86.6 | 82.5 | 241.0 | Yes |
| **YBOX** | 62.9 | 92.8 | 239.7 | 646.1 | Yes |
| **STP1** | 52.1 | 75.4 | 59.1 | 169.2 | Yes |
| **NSE** | 95.8 | 97.8 | 114.1 | 304.1 | Yes |
| **GDA** | 109.8 | 125.2 | 114.1 | 93.0 | No |
| **CRMP1** | 63.8 | 93.7 | 87.9 | 199.6 | Yes |
| **CRMP2** | 102.9 | 120.3 | 94.3 | 77.9 | No |

EU, ELISA units; SLE, systemic lupus erythematosus

In general, anti-cardiolipin antibody did not interfere significantly with the ELISAs, i.e. did not cause the recovered EU to exceed the preliminary cutoff. CRMP2 and GDA recovered EU values exceeded the preliminary cutoffs for those two ELISAs. Since these are preliminary cutoffs and significant interference was seen with IgG, a more extensive investigation of this interference will be carried out. In addition, a limitation was added to further investigate any sample that generates a positive result on all eight ELISAs.

### Establishment and Verification of Preliminary Cutoffs

A set of 200 ACD plasma samples collected from healthy female donors at the San Diego Blood Bank were used to establish preliminary thresholds for each ELISA in the development lab. The results were analyzed for normal distribution and assessed at mean +2SD, mean +3SD, and 97.5^th^ percentile as potential preliminary thresholds for positivity. Between 24 and 29 available clinical samples previously assigned positive or negative by the UC Davis feasibility assays were tested using these preliminary thresholds and plotted to visualize the discrimination between autism positive and negative samples according to the UC Davis feasibility assay. (Figure 2)

**Figure 2.**
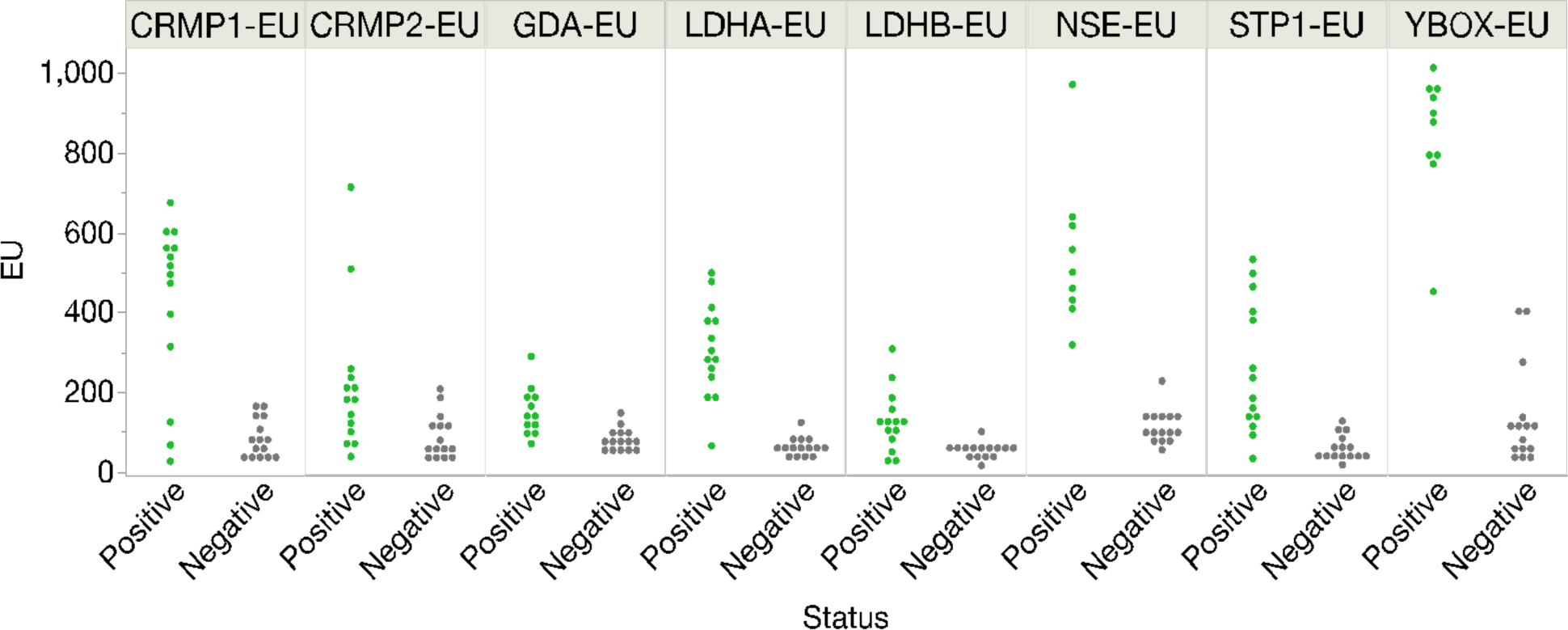
Overview of all markers by sample status. ELISA results for 24-29 clinical samples previously assigned as positive or negative. Tested using preliminary thresholds. Abbreviations: EU, ELISA units

In addition, the Youden Index (YI) for this sample set was generated using RoC analysis to help identify cutoff thresholds that best balance sensitivity and specificity. These Y1-derived thresholds were then compared to the preliminary thresholds (97.5^th^ percentile and mean +3SD) to determine which cutoff produced the optimal % percent recovery and agreement with a reference assay (the UC Davis feasibility results), as summarized in Table 12.

**Table 12.** Agreement of verification results using different thresholds.

| **ELISA** | **M+3sd** | **97.5th** | **Y1** |
| --- | --- | --- | --- |
| **Anti-CRMP1** | 89.7% | 89.7% | 89.7% |
| **Anti-GDA** | 59.3% | 59.3% | 85.2% |
| **Anti-NSE** | 100% | 100% | 100% |
| **Anti-LDHB** | 69.0% | 79.3% | 86.2% |
| **Anti-YBOX** | 96.0% | 96.0% | 100% |
| **Anti-STP1** | 79.3% | 79.3% | 86.2% |
| **Anti-CRMP2** | 72.4% | 72.4% | 75.9% |
| **Anti-LDHA** | 89.7% | 86.2% | 96.6% |

M+3sd, mean + 3 standard deviations; 97.5<sup>th</sup>, 97.5<sup>th</sup> percentile; Y1, Youden Index

Overall, the 97.5th percentile cutoff generated the best agreement metrics with the feasibility study results for the analytes in the verification dataset. However, in some cases, the YI improved agreement, indicating that the preliminary cutoffs could be further optimized. High agreement with the feasibility assay (>70%) was not always anticipated. This is because the verification assay included substantive improvements, such as different protein expression methods and optimized parameters and components.

These cutoffs are preliminary in nature and will be updated based on the results of the clinical validity study. NSE and YBOX demonstrated the best separation at the 97.5^th^ threshold, and CRMP1, LDHA, LDHB & STP1 also demonstrated acceptable separation at these thresholds. Lowering the threshold for GDA to the YI value improved the separation. Separation was worst for CRMP2 and did not show improvement using the YI.

### CLIA Validation

The ELISAs were validated in the CLIA lab to demonstrate that the performance observed during evaluation could be repeated. Results for all of the ELISAs met the acceptance criteria for linearity, precision, accuracy, analytical specificity, analytical sensitivity, sample shipping and storage, stability and lot-to-lot comparison. (Data not shown)

## DISCUSSION

The identification of specific combinations of autoantibodies targeting proteins highly expressed in the developing brain by the Van de Water laboratory, supported by a series of independent epidemiological studies as well as the critical animal models pointing to the pathogenicity of these antibodies, are important steps in defining a subset of autism with an immune-mediated etiology.^4–11,16–24^ Further, such identification of these mechanistic biomarkers highlights the importance of rigorous translation into the clinical laboratory, where they may enable earlier diagnosis and inform potential therapeutic or preventative strategies for this subset of autism. The analytical validation of these qualitative indirect ELISA IgG autoantibodies for use as a robust and reproducible highly predictive clinical grade test is a critical first step in translating these academic discoveries into a CLIA laboratory. The importance of rigorous analytical validation of these eight ELISAs is accentuated by the specific combinatorial relationship to clinical outcomes.

The clinical test for MARA discussed herein, based on the presence of one or more analytically validated autoantibody combinations targeting eight proteins critical to the developing brain, is intended to provide information as to whether a child or intended offspring is in fact at a very high likelihood of autism, compared to their already elevated risk based on their early clinical signs or a mother having had a prior autistic child. Most experts believe that early intervention in autistic children has a significant impact on subsequent development.^45–51^ Therefore, the robustness of the MAR-Autism^TM^ test, in terms of providing a quantifiable early risk indication, is hypothesized to shorten the timeline to appropriate treatment, and could thus significantly improve autism outcome. Mothers with an existing autistic child who are interested in another pregnancy may also benefit from the test for pre-pregnancy risk assessment and decision-making since a positive result in the context of a previously affected child would suggest a recurrence risk much higher than the current expectation of 19-20% based on the prior child’s affirmative diagnosis of autism.^52^ Informed decision making prior to conception of another pregnancy is vital in such settings as many families simply opt to stop reproducing after an affected child.^53^ However, it is vital to emphasize that the test is not validated for the evaluation of an ongoing pregnancy.

The pathogenic mechanism whereby maternal auto/allo-antibodies cross the placenta and target related fetal and/or newborn antigens causing disease is well known, such as in lupus related congenital heart block^54^, neonatal myasthenia gravis^55,56^, and hemolytic disease of the fetus and newborn.^57,58^ As in the setting of disorders due to other maternally transmitted autoantibodies, new strategies to remove such autoantibodies, block their transmission or otherwise mitigate their impact are now a reality. Several approaches, similar to those emerging as treatment of more well-known neonatal autoimmune disorders, are potentially relevant and are being evaluated to move the MAR-Autism^TM^ test forward as a companion diagnostic. The MAR-Autism^TM^ test has the potential to serve as a companion diagnostic for interventions such as IVIG, a biologic consisting of pooled antibodies from multiple unrelated individuals used to manage various immunodeficiency states and a variety of other conditions, including infectious, autoimmune, and inflammatory states, however the exact mechanism of action for IVIG is unclear.^59^ One mechanism may be that IVIG saturates the receptor (FcRn) responsible for transfer of maternal immunoglobulins across the placenta to the fetus, potentially reducing fetal exposure to pathogenic antibodies.^59–68^ It has been broadly and safely used with varying benefits in relevant settings, including prenatally for primary infertility and recurrent pregnancy loss, and in autism in the context of antibodies not maternally transferred but present in some older affected children through other mechanisms.^59–68^

There is an emerging broad class of monoclonal antibodies targeting FcRn, which in addition to facilitating a rapid elimination of the pathogenic endogenous IgGs, can prevent the transfer of maternal alloantibodies to the fetus.^68,69^ Several are already in use in autoimmune disorders, such as myasthenia gravis (MG)^56^ and more are in clinical trials. including in pregnancy for Hemolytic Disease of the Fetus and Newborn (HDFN) now in a Phase III trial^57,58^ where it appears more effective than IVIG. Additional strategies to lower or eliminate maternal autoantibodies related to autism include other classes of emerging agents which can selectively target and degrade these autoantibodies.^68,69^ This approach has been piloted in our animal model, which demonstrated the ability to lower one of our specific antibodies used in the MAR-Autism^TM^ test. Further evaluation of these and other similar approaches are currently underway.^71,72^

ELISA, the clinical workhorse for assessment of antibodies, is prone to imprecision for a variety of reasons and analytical error can be reduced by the validation strategies employed in this study. The importance of verifying the analytical performance of each individual antibody assay in the clinical laboratory, as described herein, cannot be understated given their role in the potential diagnosis of this subset of autism due to specific combinations of maternal autoantibodies transmitted during pregnancy. The predictive value of the autoantibody combinations in the MAR-Autism^TM^ test for autism prediction will be subsequently confirmed using these now validated component assays in a large clinical validity study.

## Data Availability

All data produced in the present study are either contained in the manuscript or available upon reasonable request to the authors

## Acknowledgments

The authors would like to thank: MARAbio’s initial investors including parents of children on the autism spectrum who sought to understand the biological cause of their child’s autism, participants in the UC Davis CHARGE and MARBLES studies, and Dana Barberio of Edge Bioscience Communications for her contribution in editing the manuscript.

## Abbreviations

ELISA: Enzyme-linked immunosorbent assay
LDH-A: lactate dehydrogenase A
LDH-B: lactate dehydrogenase B
GDA: Guanine Deaminase or cytpin
STIP1: stress-inducible protein
NSE: neuron-specific enolase
YBOX: Y-box-binding protein 1
CRMP1: collapsin response mediator protein 1
CRMP2: collapsin response mediator protein 2
LOB: Limit of Blank
LOD: Limit of Detection
LOQ: Lower Limit of Quantification
CI: confidence interval
ACD: acid citrate dextrose
RF: rheumatoid factor
SLE: systemic lupus erythematosus
YI: Youden’s index
OD: optical density or absorbance
FcRn: neonatal fragment crystallizable (Fc)-receptors
RoC: Receiver operator curve
LDT: Laboratory Developed Test

